# Validation of a Derived International Patient Severity Algorithm to Support COVID-19 Analytics from Electronic Health Record Data

**DOI:** 10.1101/2020.10.13.20201855

**Authors:** Jeffrey G Klann, Griffin M Weber, Hossein Estiri, Bertrand Moal, Paul Avillach, Chuan Hong, Victor Castro, Thomas Maulhardt, Amelia LM Tan, Alon Geva, Brett K Beaulieu-Jones, Alberto Malovini, Andrew M South, Shyam Visweswaran, Gilbert S Omenn, Kee Yuan Ngiam, Kenneth D Mandl, Martin Boeker, Karen L Olson, Danielle L Mowery, Michele Morris, Robert W Follett, David A Hanauer, Riccardo Bellazzi, Jason H Moore, Ne-Hooi Will Loh, Douglas S. Bell, Kavishwar B Wagholikar, Luca Chiovato, Valentina Tibollo, Siegbert Rieg, Anthony L.L.J. Li, Vianney Jouhet, Emily Schriver, Malarkodi J Samayamuthu, Zongqi Xia, The Consortium for Clinical Characterization of COVID-19 by EHR (4CE) (CONSORTIA AUTHOR), Isaac S Kohane, Gabriel A Brat, Shawn N Murphy

## Abstract

**Introduction:** The Consortium for Clinical Characterization of COVID-19 by EHR (4CE) includes hundreds of hospitals internationally using a federated computational approach to COVID-19 research using the EHR.

**Objective:** We sought to develop and validate a standard definition of COVID-19 severity from readily accessible EHR data across the Consortium.

**Methods:** We developed an EHR-based severity algorithm and validated it on patient hospitalization data from 12 4CE clinical sites against the outcomes of ICU admission and/or death. We also used a machine learning approach to compare selected predictors of severity to the 4CE algorithm at one site.

**Results:** The 4CE severity algorithm performed with pooled sensitivity of 0.73 and specificity 0.83 for the combined outcome of ICU admission and/or death. The sensitivity of single code categories for acuity were unacceptably inaccurate - varying by up to 0.65 across sites. A multivariate machine learning approach identified codes resulting in mean AUC 0.956 (95% CI: 0.952, 0.959) compared to 0.903 (95% CI: 0.886, 0.921) using expert-derived codes. Billing codes were poor proxies of ICU admission, with 49% precision and recall compared against chart review at one partner institution.

**Discussion:** We developed a proxy measure of severity that proved resilient to coding variability internationally by using a set of 6 code classes. In contrast, machine-learning approaches may tend to overfit hospital-specific orders. Manual chart review revealed discrepancies even in the gold standard outcomes, possibly due to pandemic conditions.

**Conclusion:** We developed an EHR-based algorithm for COVID-19 severity and validated it at 12 international sites.

## Background and Significance

The coronavirus disease 2019 (COVID-19) pandemic has stretched healthcare systems around the world to capacity. The need for actionable and reliable data has highlighted the value of the Electronic Health Record (EHR). In particular, practice patterns and patient outcomes recorded in the EHR can be rapidly aggregated and analyzed to promote learning and discovery and clinical feedback. [1] Large international investments to build such research networks [2–4] have been a largely unrealized vision [5]; COVID-19 has challenged our informatics infrastructures and highlighted continued challenges and weaknesses.

The Consortium for Clinical Characterization of COVID-19 by EHR (4CE) is a recently-convened volunteer consortium of over 300 worldwide sites who are leveraging EHR and clinical expertise to develop robust informatics-driven investigations into COVID-19. The approach relies on shared analytics scripts supporting two common research analytics formats where analysis is local, and aggregation is central. [6,7] By leveraging the investment in standard analytic models while respecting the issue of governance, we completed the initial phase of the study within two months of the COVID-19 outbreak, wherein we characterized COVID-19 comorbidities and laboratory test values at 96 international hospitals. [8]

To understand patient disease courses and investigate outcomes using EHR data, it is critical to have a reliable and robust measure of disease severity. Intuitively, outcomes such as ICU admission and death would be good correlates of severity. Early work in 4CE attempted to use these outcomes as severity measures, but it became apparent that these data are not reliably available in all environments. Therefore, 4CE sought to develop a reasonable proxy measure of worse outcomes in hospitalized patients with COVID-19 based on widely available EHR data (e.g., medication codes, diagnosis codes, and labs). This algorithm is essentially a computable phenotype, which is commonly used in medical informatics to detect the presence of a disease state through proxy measures, when no single validated data element for a disease exists. Computable phenotypes are very common [9–11] because reliable mentions of diseases are rare in the clinical record, and individual diagnosis codes are mediocre predictors of the actual presence of a disease. [12]

Computable phenotypes are often defined using clinical and informatics expertise, wherein terms are specified that correlate per clinical experience with the phenotype. However, a phenotype can make sense clinically yet have poor performance due to coding anomalies and variation between sites. It is alternately possible to define phenotypes using a data-driven approach that uses statistical algorithms to find predictors of the desired outcomes directly from the data. These can also exhibit generalization problems due to overfitting. Thus, an important next step for either approach is to validate the phenotype, which can be done by comparing the concordance between the derived phenotype and the desired outcome - for which it is a proxy - at multiple sites. Although a variety of methods for defining an outcome are possible, the most reliable method of validating a computable phenotype is to perform chart review, which is considered the best available source of truth about the patient. [13,14] For example, identification of ICU admissions might not always be accurate, especially in a pandemic situation like COVID-19, where formal protocols were not followed. In hospitals where hallways were converted into ad-hoc ICUs to support the surge of sick patients, standardized EHR data elements such as ‘transfer to ICU’ would not be properly recorded. Manual note review (and perhaps NLP in the near future) would be the only method to discover a patient’s ICU status.

### Existing Severity Measures

There has been heightened interest in disease severity measures since the outbreak of COVID-19 [15] We performed a review of 26 early COVID-19 studies. Five used ICU admission as the severity measure, one used American Thoracic Society criteria for severity of community-acquired pneumonia [16], and all the rest used the World Health Organization definition [17]. Other severity measures have been suggested [18]; however, they are not widely used or well validated.

The WHO broadly defines “severe” disease as fever or suspected respiratory infection, plus one of the following: respiratory rate >30 breaths/min, severe respiratory distress, or arterial oxygen saturation measured by pulse oximeter (SpO2) ≤93% while breathing room air. [17] The WHO definition includes patients admitted to the hospital with pneumonia who can be managed on medical wards and are not critically ill. Best evidence suggests that about 85% of such patients will never progress to critical illness such as Acute Respiratory Distress Syndrome (ARDS). [19]

ICU admission is not a useful severity measure for 4CE because these data have limited availability in EHRs. 4CE is only collecting common EHR data classes (demographics, diagnoses, medications, labs, and ICD procedure codes) and thus a severity measure must include only these. The WHO definition has the same issue and is also very inclusive. It is most strongly a proxy for hospital admission (moderate disease) rather than a difficult hospital course (severe disease). As such, it is a too sensitive marker for 4CE’s goal of identifying severe patients.

### Objective

We set out to develop an EHR-driven severity algorithm as a proxy for worse clinical course in hospitalized patients with COVID-19 and validate this computable phenotype algorithm against the outcomes of ICU admission and/or death. Because outcome data had uncertain accuracy, we performed a limited chart review to better understand validation performance. Finally, we compared a data-driven algorithm at one site to the expert-derived 4CE algorithm.

## Materials and Methods

### Defining Severity

First, we developed a 4CE severity algorithm that is both clinically reasonable and possible to identify at our diverse sites. To do this, we needed to limit severity to the EHR data classes that 4CE is collecting: demographics, diagnoses, medications, labs, and ICD procedure codes. We did not use outcomes (e.g., ICU admission), symptoms (e.g., wheezing), or vital signs (e.g., respiratory rate), as these are not widely or reliably available in electronic health records.

We used the WHO severity definition as a starting point and two authors (GW and GB) identified a much more specific diagnosis group: patients who required invasive mechanical ventilation for acute respiratory failure or vasoactive medication infusions for shock.

We created a value-set of EHR data elements that suggest these disease states, based on commonly available data classes:

- **Lab Test:** PaCO2 or PaO2
- **Medication:** sedatives/anesthetics or treatment for shock
- **Diagnosis:** ARDS, ventilator-associated pneumonia
- **Procedure:** endotracheal tube insertion or invasive mechanical ventilation [20]

These data elements correlate with many individual standard codes. To identify standard codes, we cross-referenced the i2b2 ontology in the ACT network. [2] This is a comprehensive terminology dictionary of 2.5 million codes found in many EHRs, arranged hierarchically so that individual codes are arranged in folders describing the above concepts. The result was a list of ∼100 codes in the International Classification of Diseases versions 9 and 10 (ICD-9 and ICD-10), Logical Observation Identifiers Names and Codes (LOINC), and RxNorm format, which are international standards used for research. These are listed in Table A1 in the Appendix.

Local sites expanded these standard codes to match their local codes. Often, this was assisted with previous mappings from i2b2 where local items were a child folder of the standard code. [21] When mappings were not straightforward, the terms that most closely matched the definition were used, maximizing semantic equivalence across sites. For example, some US sites had both CPT and ICD procedure codes; the CPT codes were not added when ICD was available. In contrast, because some European sites do not use ICD-10-CM, other coding systems (like OPS codes) were added to identify invasive mechanical ventilation.

Each participating site defined a “4CE Cohort” as any hospitalized patient who had any positive test for SARS-CoV-2 +14/-7 days around the hospitalization. A patient was considered severe if codes generated during the hospital course matched any code in the value set.

### Network-Wide Analysis: 4CE Severity Validation

To validate the 4CE severity algorithm, a subset of sites identified patients who were admitted to the ICU and/or who died. Although these outcomes were not a perfect equivalence to severe disease or hospital course, they are objective measures that can be gleaned from patient data. We defined three options for confirming ICU admission, in order from most to least accurate:

1. **Chart Review**. This is considered the gold-standard for identifying outcomes like ICU admission and could have been particularly useful in crisis situations like the COVID-19 pandemic. Nonetheless, chart review is time-consuming and laborious, so this option was impractical at institutions without substantial human resources.
2. **Local Hospital Data**. Hospital systems have idiosyncratic methods of determining ICU status, but they tend to be fairly accurate because they are used to determine admission, discharge, and transfer (ADT) status and to manage hospital bed allocation. However, not all sites had access to local hospital data, and expertise was required to incorporate this information into a data warehouse. Such limitations underscored the rationale for development of the severity proxy.
3. **Specific ICU CPT Procedure Codes**. In the US, healthcare providers and hospitals use current procedural terminology (CPT) codes to bill for provided critical care services. CPT codes for billing time spent providing critical care (99291, 99292) provide a third option for defining ICU admission. These CPT codes were not used to define the severity proxy measure.

Each site computed a set of 2×2 tables comparing the 4CE severity algorithm to three outcomes (death only, ICU only, and ICU-or-death) (Table A2). Sites calculated sensitivity, specificity, positive predictive value (PPV), negative predictive value (NPV), and F1-score from these tables. We used a fixed-effects meta-analysis model to estimate pooled scores over all sites. Sites then calculated the performance of individual code classes by computing the sensitivity for the same set of three outcomes. This analysis gave further insight into the components of the algorithm’s performance at each site. Sensitivity would be highest for the full algorithm, as it is a combination of all 4CE severity code classes. Additionally, each site reported its approach for confirming ICU admission, total number of ICU beds (to give a sense of site capacity), and any variation from the standard 4CE severity definition or cohort definition. Sites performed these analyses between August 5, 2020 and September 18, 2020, reflecting cases that were recorded from March through August 2020.

To understand the practical differences between methods of defining ICU admission, we performed a limited analysis at two sites. We used a set of chart-reviewed ICU admission data among 866 confirmed COVID-19 patients from Massachusetts General Hospital (MGH) between March 8, 2020 and June 3, 2020. Extensive manual chart reviews were completed by trained reviewers, including physicians, pharmacists, research nurses, and clinical research coordinators. [22] University of Freiburg Medical Center in Germany (UKFR) provided a set of ICU admission flags obtained from manual chart review of 168 patients in their 4CE COVID-19 cohort. They identified ICU stays directly related to their COVID-19 infection. We compared coded ICU admissions to the chart-reviewed data at MGH and UKFR for patients in the 4CE COVID-19 cohort. These overlapping data sets allowed us to compare the two definitions of ICU admission with the 4CE severity algorithm. We also compared the performance of the chart-reviewed definition to CPT code-based ICU admission (99291 and 99292) using MGH data.

### Data-Driven Pilot Analysis

We undertook a machine-learning approach at a single site, Mass General Brigham, using an existing computable phenotyping pipeline.

First, we evaluated the classification performance of the 4CE severity algorithm. Second, we performed automated computable phenotyping using the Minimize Sparsity Maximize Relevance dimensionality reduction algorithm to select codes from among all possible data elements. [23,24] In both approaches, we applied generalized linear models (GLM) with a logit link, binomial distribution, and component-wise functional gradient boosting [25,26] to develop the computational models. We used the 4CE COVID-19 cohort with ICU admission and/or death as the target for prediction. We trained and tested the models using an 80-20 train-test split, which we iterated 9 times to capture potential variability in performance metrics due to sampling. Model tuning was performed via 5-fold cross-validation. To evaluate the two computable phenotyping models, we calculated the area under the receiver operating characteristic curve (AUC ROC) on the held-out test sets.

## Results

### 4CE Severity Analysis

Twelve sites participated in this analysis. The site names, locations, number of hospital beds, number of ICU beds (not reflecting surge capacity), and total 4CE cohort size (rounded to the nearest 10) are shown in Table 1. We also included the data source used for ICU admission and whether the site’s code mapping included any significant additions to the severity value set. (For example, European sites do not use the US Clinical Modifications of ICD-10, so additional standard codes were needed.) In further results, site names were replaced by a randomly-assigned region identifier (either USAx for sites in the United States or GLOBALx for others).

**Table 1:** Participating 4CE sites and metadata on ICU and 4CE coding definitions, number of beds, and 4CE cohort size (rounded to the nearest 10).

| Healthcare System | City | Country | # Hospitals | Total Beds | ICU Beds | ICU Data Source | 4CE Cohort Size | Additional Codes in 4CE Severity Value Set |
| --- | --- | --- | --- | --- | --- | --- | --- | --- |
| Mass General Brigham (Partners Healthcare) | Boston, MA | USA | 10 | 3418 | 292 | hospital data | 3290 | None |
| University of Pennsylvania | Philadelphia, PA | USA | 5 | 2469 | 515 | hospital data | 2330 | Hospital Data for Intubation and Ventilation |
| University of Pittsburgh | Pittsburgh, PA | USA | 39 | 8400 | 589 | CPT code and hospital location | 990 | CPT Codes for Intubation and Ventilation |
| Beth Israel Deaconess Medical Center | Boston, MA | USA | 1 | 673 | 77 | hospital data | 690 | None |
| University of Michigan | Ann Arbor, MI | USA | 3 | 1043 | 141 | CPT code and hospital location | 420 | None |
| University of California, Los Angeles | Los Angeles, CA | USA | 2 | 786 | 192 | hospital data | 430 | None |
| Bordeaux University Hospital | Bordeaux | France | 3 | 2676 | 180 | hospital data | 360 | CCAM (French procedure codes) |
| Istituti Clinici Scientifici Maugeri | Pavia, Lumezzane/Brescia, Milan | Italy | 3 | 775 | 0 | N/A (Rehab hospital - no ICU) | 260 | None |
| Medical Center, University of Freiburg | Freiburg | Germany | 1 | 1660 | 132 | hospital data | 190 | ICD-10 GM and OPS codes |
| Boston Children's Hospital | Boston, MA | USA | 1 | 404 | 107 | ICU note type | 60 | None |
| National University Hospital | Singapore | Singapore | 1 | 1556 | 65 | hospital data | 260 | SNOMED codes for diagnoses; TOSP billing codes for procedures |
| St. Luke's University Health Network | Bethlehem, PA | USA | 12 | 1700 | 287 | hospital data | 1230 | None |

Sites reported the sensitivity, specificity, PPV, and NPV of the 4CE severity algorithm for the outcome of ICU admission and/or death. The pooled F-score over 12 sites was estimated as 0.72 (95% CI: 0.63,0.80) using a fixed-effect meta-analysis model. The pooled sensitivity was 0.73 (95% CI: 0.64,0.82) with mean 0.73 (range 0.56). The pooled specificity was 0.83 (95% CI: 0.76,0.91) with mean 0.80 (range 0.5). The sensitivity, specificity, PPV, NPV, and F-score by site can be seen in Table 2. Sites also computed these measures separately for ICU admission and death. The pooled specificity went down for the individual outcomes (0.79 for ICU and 0.67 for death), but sensitivity was higher (0.77 for ICU, 0.76 for death). The statistics for the individual outcomes can be seen in Table A3 in the Appendix.

**Table 2.**
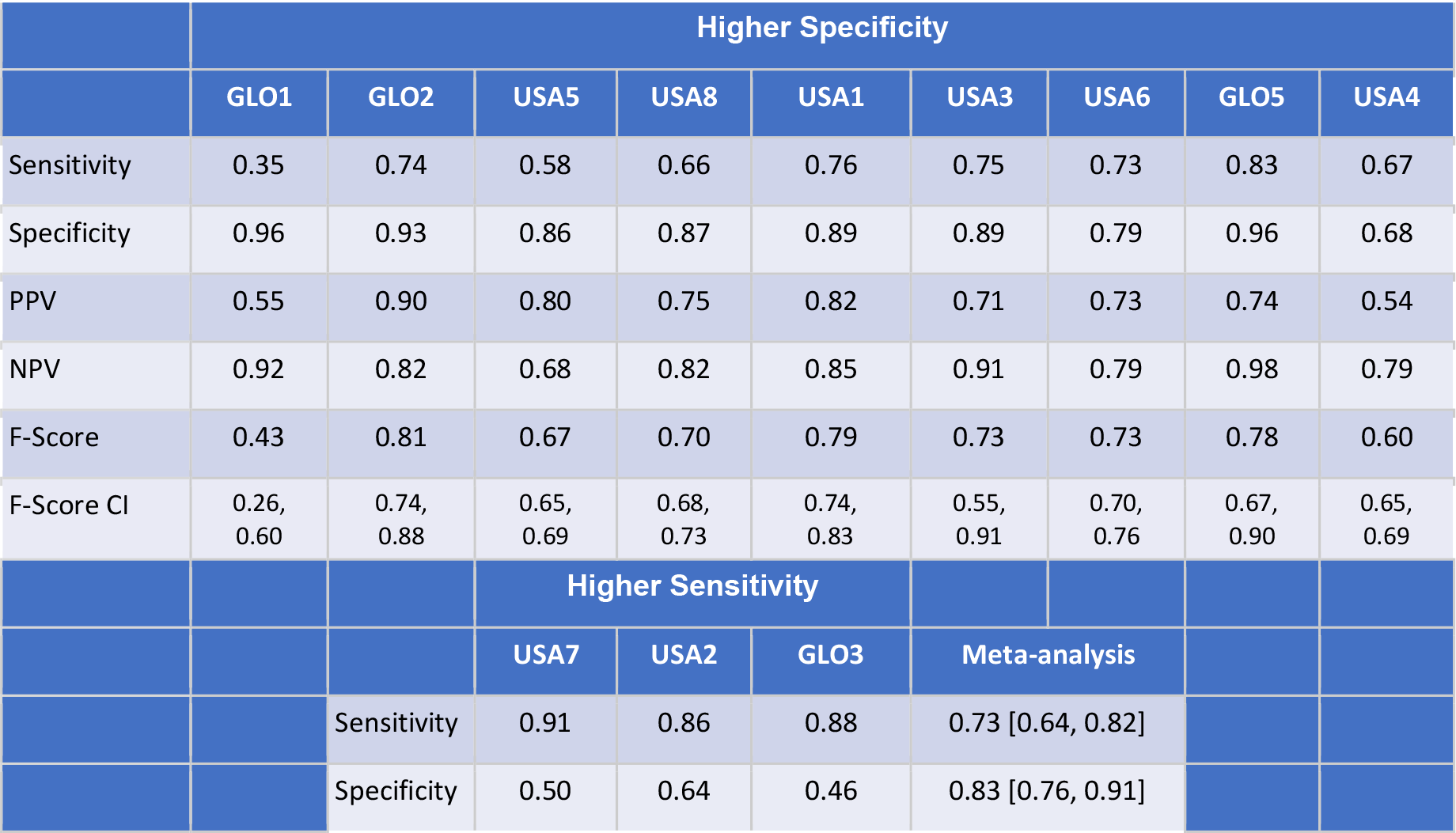
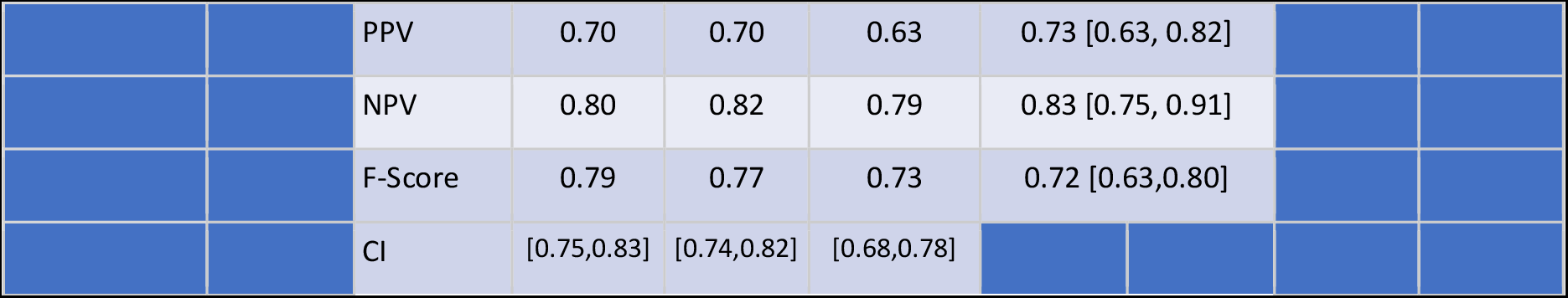
The sensitivity, specificity, PPV, NPV, and F1-score of the 4CE severity algorithm for the outcome ICU admission and/or death, at each site in the United States (USA) and outside the US (GLObal). Estimates of the pooled scores were computed using a fixed-effect meta-analysis model.

|  | Higher Specificity |  |  |  |  |  |  |  |  |
| --- | --- | --- | --- | --- | --- | --- | --- | --- | --- |
|  | GLO1 | GLO2 | USA5 | USA8 | USA1 | USA3 | USA6 | GLO5 | USA4 |
| Sensitivity | 0.35 | 0.74 | 0.58 | 0.66 | 0.76 | 0.75 | 0.73 | 0.83 | 0.67 |
| Specificity | 0.96 | 0.93 | 0.86 | 0.87 | 0.89 | 0.89 | 0.79 | 0.96 | 0.68 |
| PPV | 0.55 | 0.90 | 0.80 | 0.75 | 0.82 | 0.71 | 0.73 | 0.74 | 0.54 |
| NPV | 0.92 | 0.82 | 0.68 | 0.82 | 0.85 | 0.91 | 0.79 | 0.98 | 0.79 |
| F-Score | 0.43 | 0.81 | 0.67 | 0.70 | 0.79 | 0.73 | 0.73 | 0.78 | 0.60 |
| F-Score CI | 0.26, 0.60 | 0.74, 0.88 | 0.65, 0.69 | 0.68, 0.73 | 0.74, 0.83 | 0.55, 0.91 | 0.70, 0.76 | 0.67, 0.90 | 0.65, 0.69 |
|  |  |  | Higher Sensitivity |  |  |  |  |  |  |
|  |  |  | USA7 | USA2 | GLO3 | Meta-analysis |  |  |  |
|  |  | Sensitivity | 0.91 | 0.86 | 0.88 | 0.73 [0.64, 0.82] |  |  |  |
|  |  | Specificity | 0.50 | 0.64 | 0.46 | 0.83 [0.76, 0.91] |  |  |  |

|  |  |  |  |  |  |  |
| --- | --- | --- | --- | --- | --- | --- |
|  |  | PPV | 0.70 | 0.70 | 0.63 | 0.73 [0.63, 0.82] |
|  |  | NPV | 0.80 | 0.82 | 0.79 | 0.83 [0.75, 0.91] |
|  |  | F-Score | 0.79 | 0.77 | 0.73 | 0.72 [0.63,0.80] |
|  |  | CI | [0.75,0.83] | [0.74,0.82] | [0.68,0.78] |  |

Sites computed the sensitivity of individual code classes to understand how each contributed to the performance of the overall metric. Code classes demonstrated high variability of sensitivity across sites (Figure 1). For example, the anesthetic medication class had sensitivity ranging from 0.02 to 0.67. Code class sensitivity for the separate outcomes of ICU admission and death can be seen in Figures A1-A3 in the Appendix. Figure 2 shows the percentage of all severe patients with a code in each class with the outcome of ICU admission and/or death. Figure 3 shows the overlap of high-level code classes in a Venn Diagram.

**Figure 1.**
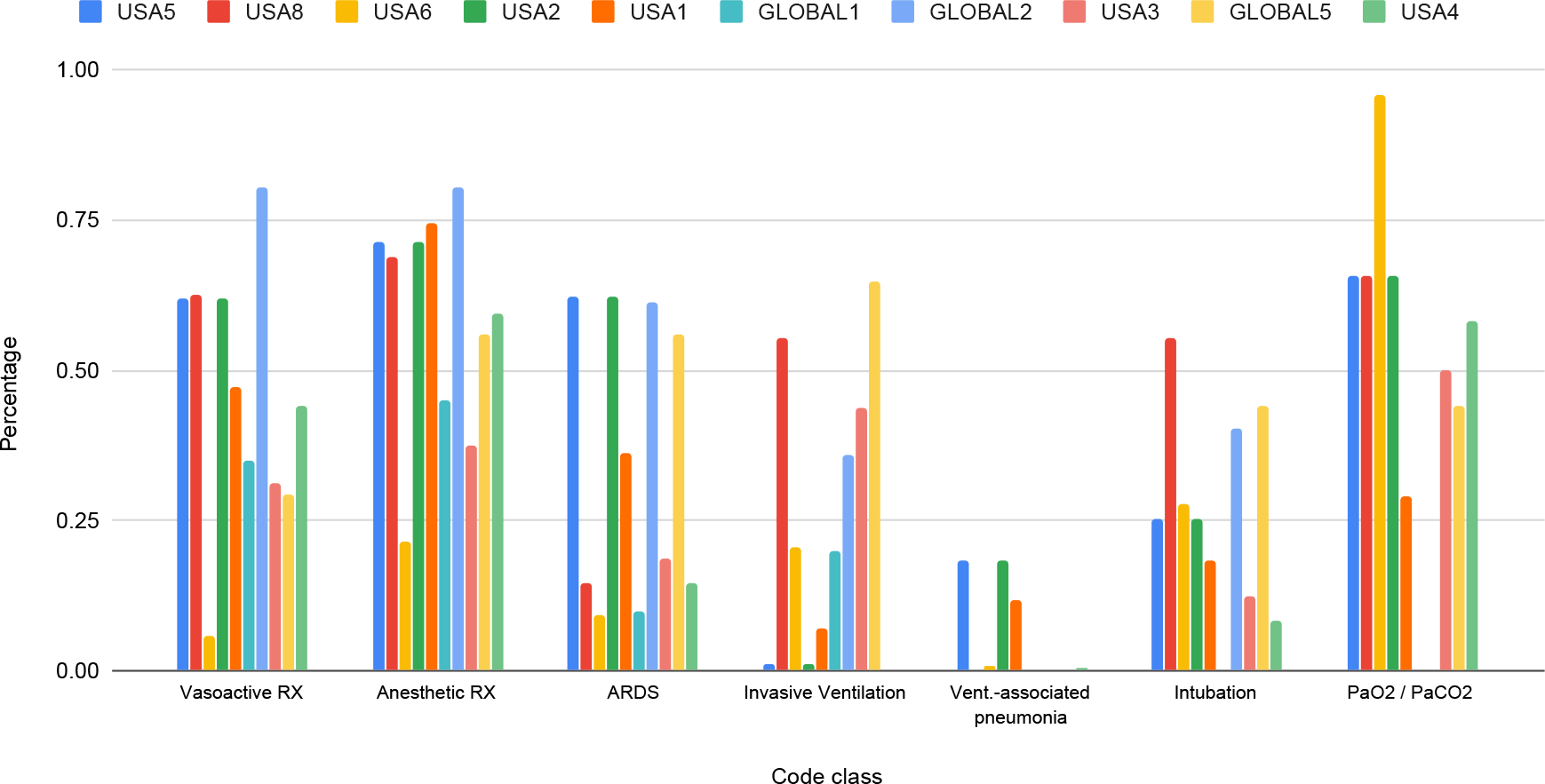
Sensitivity of code classes to identify ICU admission and/or death.

**Figure 2.**
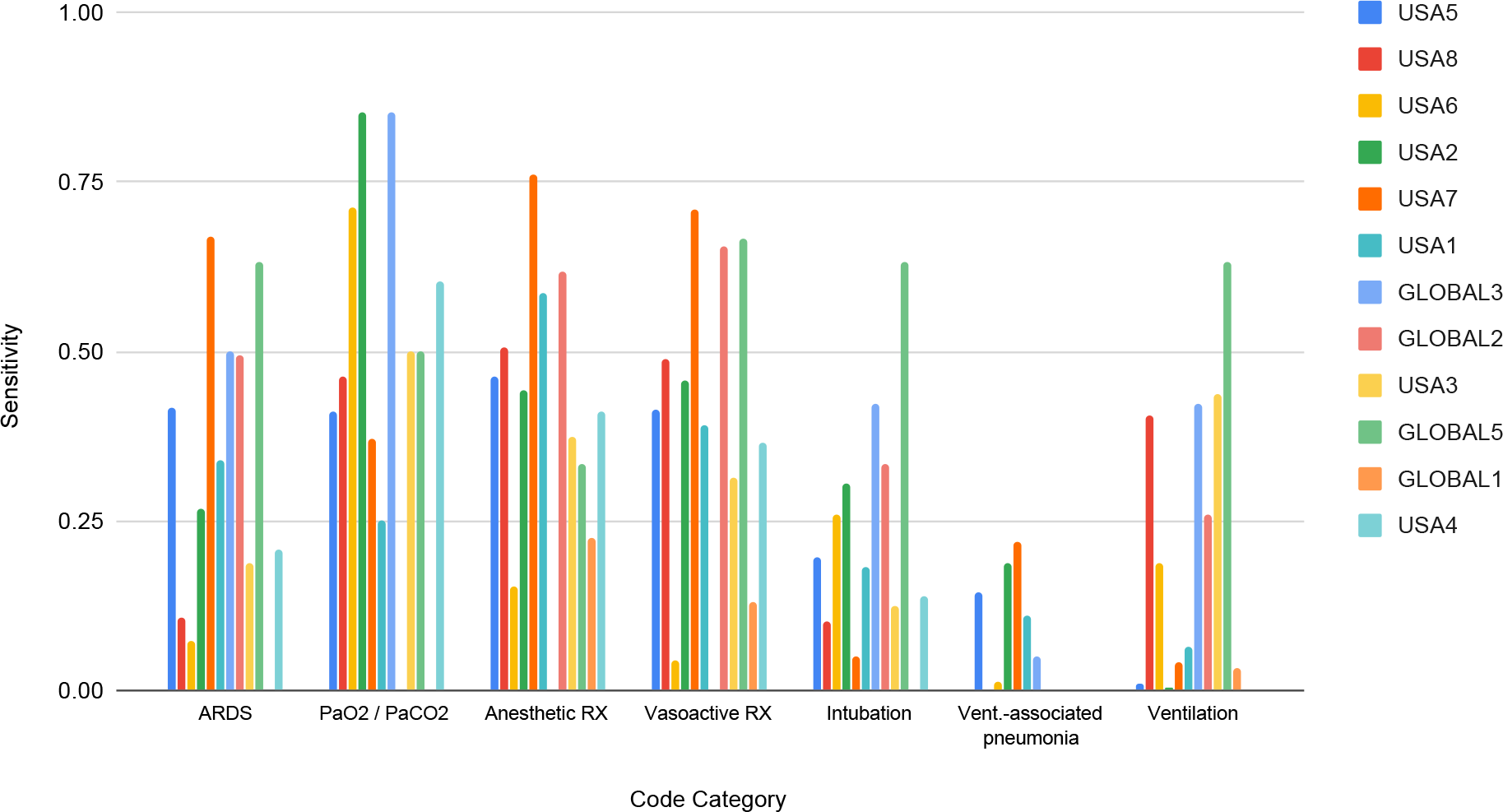
Percentage of patients identified by 4CE severity algorithm and having the outcome ICU admission and/or death (i.e., true positives), broken down by code class.

**Figure 3.**
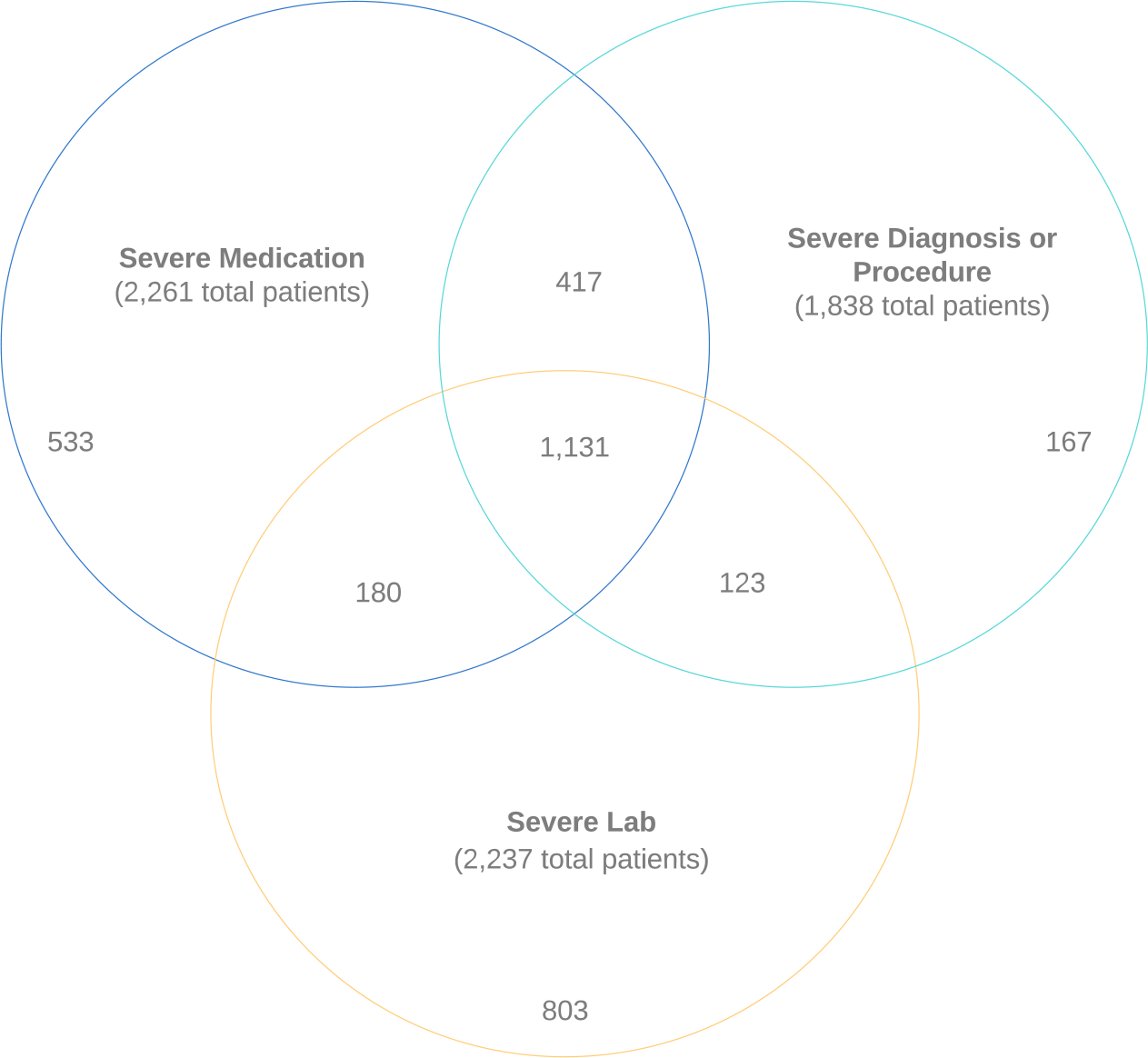
Venn Diagram showing overlap of code classes among patients with the 4CE Severe Phenotype. (Nine sites reporting).

### Comparison of ICU Definitions

We computed the precision and recall of code-defined ICU admission using chart review as the reference at MGH and UKFR. At MGH, we found agreement for ICU admission with 97% precision and 83% recall. At UKFR, we measured 78% precision and 85% recall. At MGH, we also compared agreement of CPT-code ICU admission definition to chart-reviewed ICU admission and found a 49% precision and 49% recall.

We also recomputed summary statistics of the performance of our 4CE severity algorithm for the outcome of ICU admission and/or death using the chart-reviewed definition of ICU. At MGH and UKFR, the sensitivity was higher using the chart-reviewed definition (MGH: 0.80 vs 0.58 using hospital codes; UKFR: 0.85 vs. 0.74 using hospital codes). Specificity went down at MGH (0.75 vs 0.86 using hospital codes), while it went up slightly at UKFR (0.96 vs 0.93 using hospital codes).

The differences between UKFR and MGH (lower agreement precision at UKFR, higher specificity performance of severity) are likely due to UKFR identifying only COVID-related ICU admissions, while MGH identified all ICU admissions during the COVID-19 hospitalization.

The full sets of summary statistics are reported in Tables 3 and A4.

**Table 3.** Comparing the performance of the 4CE Severity algorithm when using chart-reviewed ICU admission data to hospital codes, at MGH and UKFR. The hospital column is repeated from Table 2 for clarity.

|  | MGH: hospital | MGH: chart | UKFR: hospital | UKFR: chart |
| --- | --- | --- | --- | --- |
| Sensitivity | 0.58 | 0.8 | 0.74 | 0.85 |
| Specificity | 0.86 | 0.75 | 0.93 | 0.96 |
| PPV | 0.8 | 0.57 | 0.9 | 0.93 |
| NPV | 0.68 | 0.9 | 0.82 | 0.91 |

### Data-Driven Pilot

The GLM model trained using the 4CE severity codes performed with a mean AUC ROC 0.903 (0.886, 0.921) on the MGB COVID-19 cohort of 4227 patients. The GLM model trained on MSMR-selected codes (from among all possible diagnosis, medication, and LOINC codes) resulted in a mean AUC ROC of 0.956 (95% confidence interval: 0.952, 0.959). See Figure 4.

**Figure 4.**
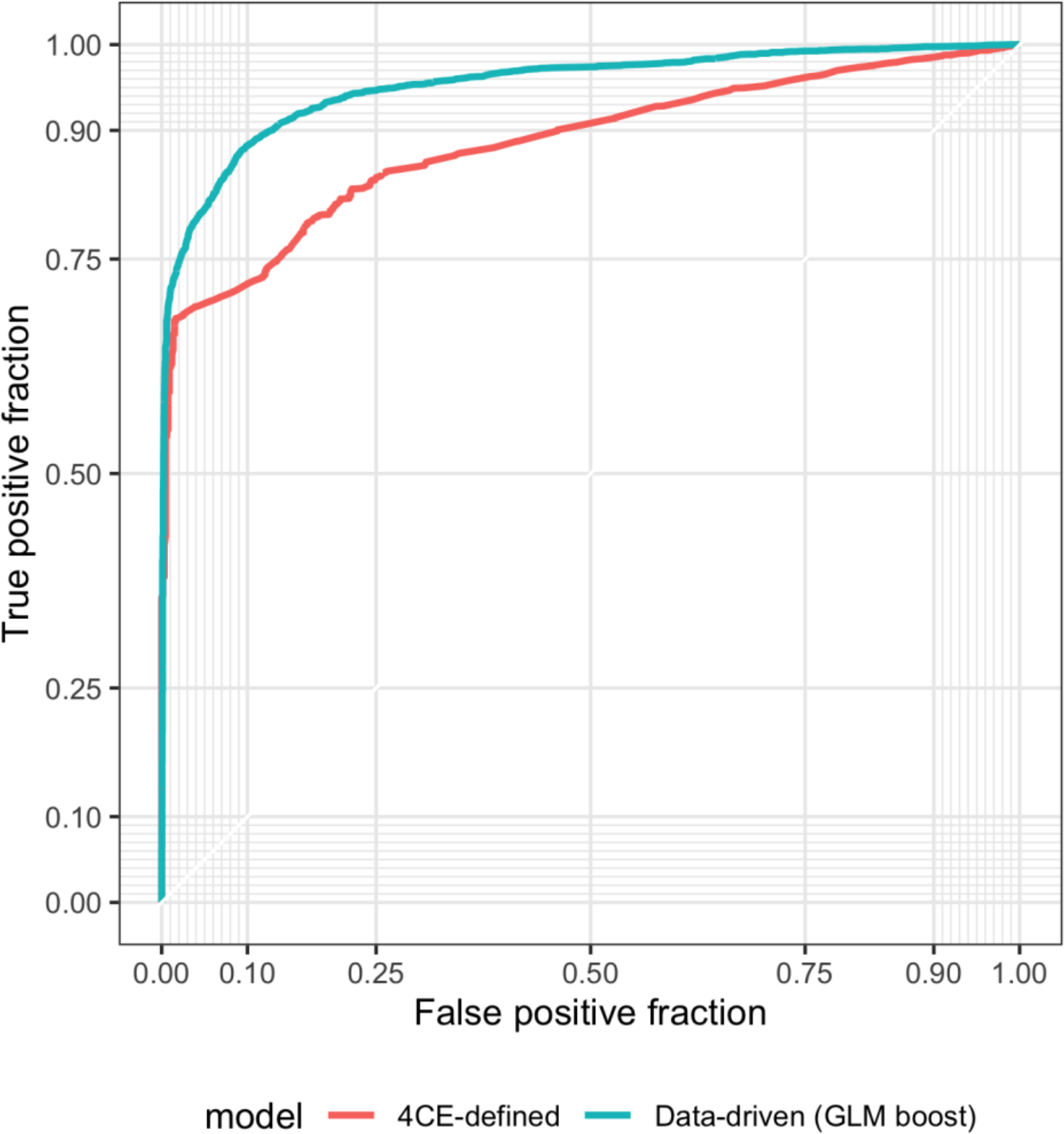
ROC curves when using a GLM boost algorithm on 4CE-defined features vs. a data-driven approach.

The MSMR-based algorithm’s top ten codes (by odds ratio) fell into the following categories:

- Similar to the 4CE definition: PaCO2, PaO2, ARDS, sedatives
- Reflective of ICU ordering patterns: d-dimer, immature granulocytes, albumin
- Surprising proxies of severity: chlorhexidine, glycopyrrolate, palliative care encounter

## Discussion

When using EHR-derived data for research, we often adopt proxies for outcomes, especially if these outcomes are infrequently or poorly recorded in the EHR. Validation of these proxies is essential so that we can understand their strengths and limitations. Furthermore, if research is to be performed on a network and especially global scale, the outcome proxies must use data types broadly available through most EHRs and also be validated at multiple sites to account for the differences of coding patterns that occur. Examining subgroup performance of the codes can further improve our ability to understand cross-site differences.

In this study, our primary aim was to develop and validate an EHR-based severity algorithm for the 4CE network to enable network-wide research on COVID-19 across numerous heterogeneous sites. The EHR proxies we used to test for severity included commonly available elements in the EHR: diagnosis codes, laboratory orders, medication orders, and procedure codes. These elements improve our ability to infer the presence of respiratory distress and shock, which presumably are serious enough to lead to ICU admission, if available, and/or death.

This study highlights the frequent presence of coding differences between sites, as demonstrated by the remarkable variation of sensitivity by code class. Moreover, the codes captured for the severity algorithm at each site are very different. For example, some sites had a very high prevalence of mechanical ventilation codes and blood gas orders, whereas others had a low prevalence of these same measures, likely due to practice variation and code extraction differences. We compensated for this limitation by employing a method that accounts for this issue and highlights the importance of expert-derived proxies for accurate EHR-based analysis. Clinicians who understood the vagaries of hospital coding helped several sites to improve their data extraction and analysis, thereby contributing to the data quality of the 4CE initiative.

Given that the codes were a proxy for the severity of illness, the PPVs we obtained in the range of 0.7 to 0.9 and the NPVs in the range of 0.68 to 0.98 are indicative of the model’s overall success. At three sites, the 4CE severity algorithm was more sensitive than specific. Patient mortality or ICU transfer was captured by the algorithm, but it also captured patients without those outcomes. At most sites (9/12), the algorithm had higher specificity than sensitivity; it flagged mostly ICU or deceased patients but missed patients as well.

This study also highlights the challenges in selecting a gold standard for validation. There is no measurable assessment of a patient’s actual complexity, so we chose ICU admission or mortality as they are commonly measured gold standards. However, ICU admission is not always clearly defined, especially during the pandemic. We evaluated 3 ways of identifying ICU admission, with accuracy improving from CPT codes to hospital code ICU designation to chart review by clinical experts. Our separate analysis of ICU admission definition suggests that the particular approach to coding ICU admission could impact measured performance. It also validated our prioritization of choices for defining ICU admission: chart review was preferred, followed by hospital codes, and then billing data. The gold standard for validation is chart review, and the differences between what is actually recorded in a patient’s chart and what data elements are available in the EHR are not always appreciated. In our analysis, chart review as compared to hospital data had precision of 97% (MGH) / 78% (UKFR) and recall of 83% (MGH) / 85% (UKFR), due largely to ICU admissions missed in hospital codes. This is probably due to the pandemic situation of COVID-19, where non-traditional spaces were converted into ICUs to support the surge of sick patients. The 4CE Severity algorithm performed overall better when using the chart review-based ICU admission numbers and was able to correctly identify many more severe patients. Sensitivity increased by 0.22 at MGH and 0.11 at UKFR. Change in specificity was mixed, but this was likely influenced by the different ICU admission targets at the two sites (all ICU admissions at MGH vs. COVID-related ICU admissions at UKFR). Billing codes were significantly less precise, missing many ICU admissions, yielding 49% precision and 49% recall. In the next phase of our work, it will be important to validate our findings with the addition of clinical notes at additional sites.

We explored a machine-learning data-driven approach at a single site and compared the results to our expert-derived algorithm. Among the top ten features identified by the data-driven model, four were conceptually similar to the expert-derived algorithm. Three were labs that occurred more frequently in the ICU than on the floor, which are reflective of biases in ordering patterns more than clinically meaningful data points. [27] The remaining orders were interesting proxies of the ICU (e.g., chlorhexidine, an antibacterial agent used for cleaning the skin). These proxies may be less generalizable than expert-curated codes.

### Limitations

Our data-driven computable exploration was only performed at one site. In the future, we hope to engage a larger sample of sites in a data-driven analysis, which would allow us to pool together a list of common codes to better discern generalizability. This will become possible as the 4CE network expands its computational infrastructure.

Additionally, all participating hospitals were in countries that experienced a surge in the COVID-19 pandemic, which could create unanticipated bias in the results.

## Conclusion

We developed an EHR-based severity algorithm that can be used when longer-term outcomes data are not readily or reliably available. We validated this at 12 international 4CE sites and confirmed its good performance, due largely to its inclusiveness and breadth. We discovered many coding differences in individual EHR elements across sites. Additionally, we explored the comparison of an expert-derived proxy to a data-driven acuity score that maximized performance at individual sites. Finally, we found differences in ICU admission definitions, so only chart review captured information that was not reliable in hospital administrative data.

## Informed Consent/IRB Statement

Each institution reported obtaining proper institutional review board approval for data sharing. Certifications of waivers or approval were collected by the consortium. See Table B2 in Appendix B for details. As data were transmitted in aggregate, no patient level data were available from any site.

## Data Availability

All data collected for this study is presented in the manuscript or appendix. The 4CE Consortium provides additional visualizations and data for other consortium projects: https://covidclinical.net

## Acknowledgements

Thanks to all members of the 4CE Consortium (see list in the Appendix) and all the effort and hard work of the local teams at the 12 sites. Thanks also to Brigitta Gough and Margaret Vella for their help in preparing this manuscript for submission, which was itself a massive undertaking.

## Contributions

Murphy, Brat, and Kohane contributed equally. Klann led the study and writing the manuscript. All authors approved the manuscript and contributed substantially. A table including full contributions is listed in Appendix B (Table B1).

## Funding

This work was supported in part by the grants in the table below.

| Grant Name | Funder | Grant # |
| --- | --- | --- |
| Integrating EHR and Genomics to Predict Multiple Sclerosis Drug Response | NIH National Institute of Neurological Disorders and Stroke | R01NS098023 |
| Pediatric Critical Care and Trauma Scientist Development Program | NIH Institute of Child Health and Human Development | K12 HD047349 |
| The Role of Angiotensin-(1-7) in Hypertension and Hypertension-Induced Heart and Kidney Damage | NIH Heart, Lung, and Blood Institute | K23-HL148394 |
| Angiotensin-(1-7) Contributes to Hypertension and Hypertension-Induced Heart and Kidney Damage | NIH Heart, Lung, and Blood Institute | L40-HL148910 |
| Instrumenting the Delivery System for a Genomics Research Information Commons | NIH National Center for Advancing Translational Sciences | U01TR002623 |
| Wake Forest Clinical and Translational Science Institute | NIH National Center for Advancing Translational Sciences | UL1-TR001420 |
| Institutional Clinical and Translational Science Award | NIH National Center for Advancing Translational Sciences | UL1-TR001878 |
| Michigan Institute for Clinical and Health Research (Michr) | NIH National Center for Advancing Translational Sciences | UL1TR002240 |
| UCLA Clinical and Translational Science Award | NIH National Center for Advancing Translational Sciences | UL1TR001881 |
| University Of Pittsburgh Clinical And Translational Science Institute | NIH National Center for Advancing Translational Sciences | UL1 TR00185705 |
| Harvard Catalyst CTSA/ACT Grant | NIH National Center for Advancing Translational Sciences | 5UL1TR001857-05 |
| Development and Evaluation of a Learning Electronic Medical Record System | NIH National Library of Medicine | R01 LM012095 |
| Biases introduced by filtering electronic health records for patients with "complete data" | NIH National Library of Medicine | R01LM013345 |
| Developing i2b2 into a Health Innovation Platform for Clinical Decision Support in the Genomics Era | NIH National Human Genome Research Institute | 5R01HG009174-04 |
| Data Fusion: A Sustainable, Scalable, Open Source Registry Advancing PVD Research | NIH National Heart, Lung, and Blood Institute | U01HL121518 |
| Medical Informatics Funding Scheme, MIRACUM Consortium | German Federal Ministry of Education and Research | FKZ 01ZZ1801B |

## Appendix A: Additional Methods and Results

### 4CE Detailed Severity Definition

The codification of the following data elements results in ∼100 codes in ICD-9, ICD-10, LOINC, and RxNorm format, international standards used for research.

- **Lab Test:** PaCO2 or PaO2
- **Medication:** sedatives/anesthetics or treatment for shock
- **Diagnosis:** ARDS, ventilator-associated pneumonia
- **Procedure:** endotracheal tube insertion or invasive mechanical ventilation These 100 elements are listed in Table A1 below.

**Table A1:**
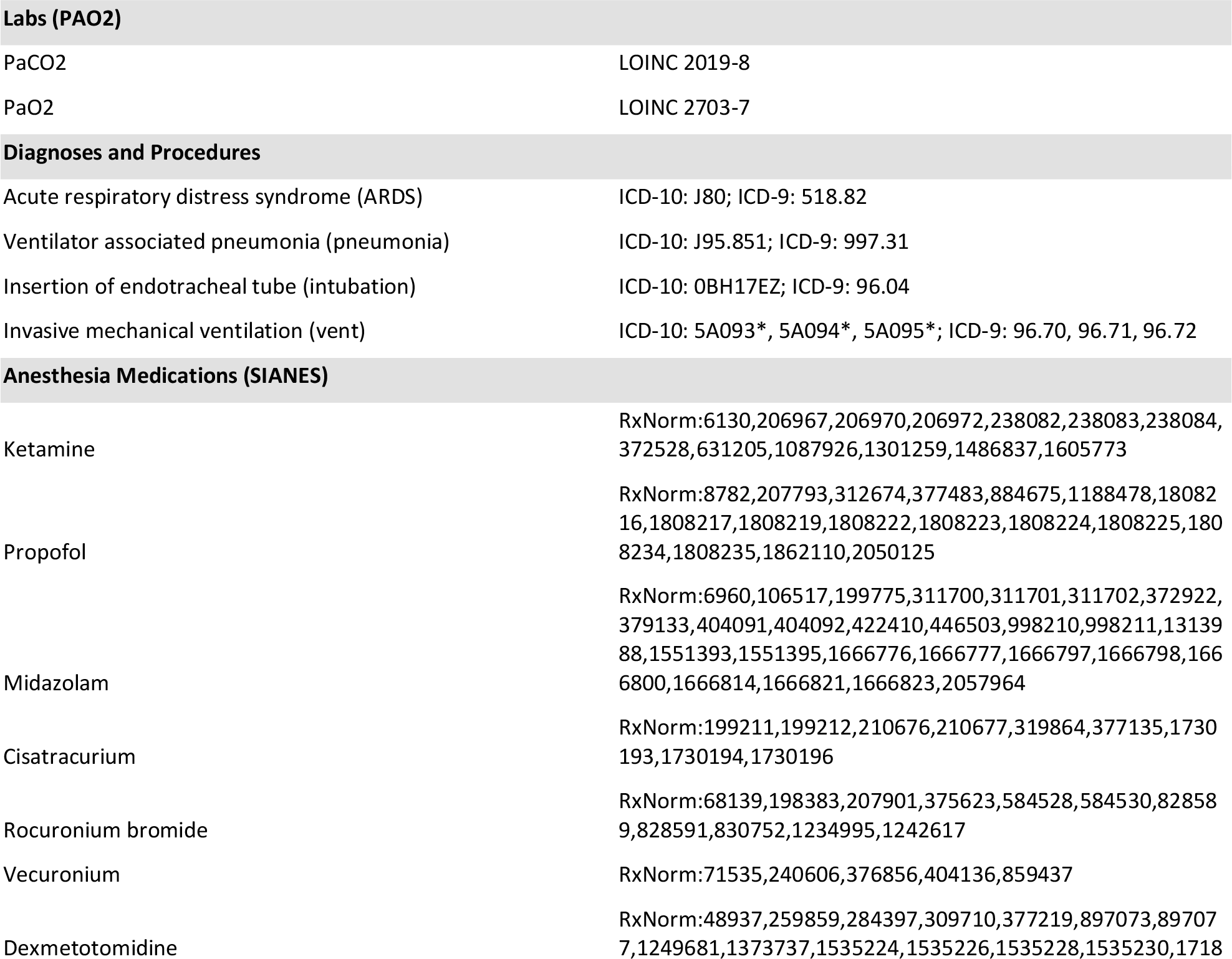
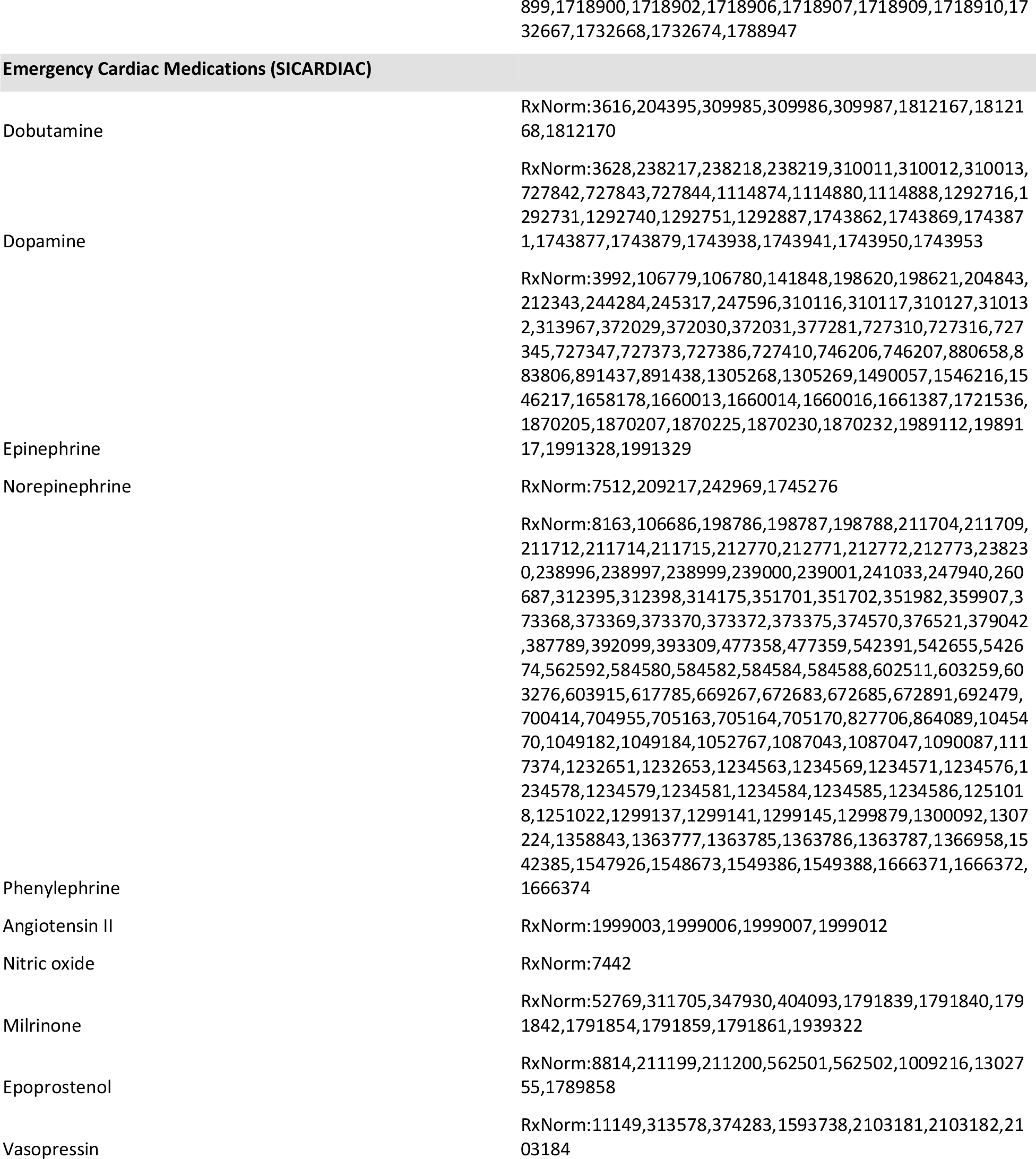
4CE Severity Codes [26].

**Table A2.** Severity Analysis 2×2 tables design.

|  | ICU and/or Death | No ICU or Death |
| --- | --- | --- |
| Severe | Severe and true | Severe only |
| Not Severe | True only | Not severe and not true |

### Network-Wide Analyses: Individual Outcomes

**Table A3.** Sensitivity, Specificity, PPV, and NPV by outcome (ICU admission and/or death, ICU, and death)

| Outcome | Measure | Meta-analyses | Mean | USA5 | USA8 | USA6 | USA2 | USA7 | USA1 | GLOB3 | GLOB1 | GLOB2 | USA3 | GLOB5 | USA4 |
| --- | --- | --- | --- | --- | --- | --- | --- | --- | --- | --- | --- | --- | --- | --- | --- |
| ICU/DEATH | Sensitivity | 0.73<br>[0.64, 0.82] | 0.73 | 0.58 | 0.66 | 0.73 | 0.86 | 0.91 | 0.76 | 0.88 | 0.35 | 0.74 | 0.75 | 0.83 | 0.67 |
| ICU | Sensitivity | 0.77<br>[0.68, 0.87] | 0.79 | 0.62 | 0.75 | 0.74 | 0.88 | 0.91 | 0.78 | 0.89 | n/a | 0.81 | 0.75 | 0.83 | 0.71 |
| DEATH | Sensitivity | 0.76<br>[0.64, 0.87] | 0.78 | 0.59 | 0.66 | 0.78 | 0.91 | 0.90 | 0.80 | 0.91 | 0.35 | 0.76 | 1.00 | 1.00 | 0.73 |
| ICU/DEATH | Specificity | 0.83<br>[0.76, 0.91] | 0.79 | 0.86 | 0.87 | 0.79 | 0.64 | 0.50 | 0.89 | 0.46 | 0.96 | 0.93 | 0.89 | 0.96 | 0.68 |
| ICU | Specificity | 0.79<br>[0.71, 0.87] | 0.75 | 0.85 | 0.85 | 0.78 | 0.62 | 0.45 | 0.88 | 0.41 | n/a | 0.89 | 0.89 | 0.96 | 0.67 |
| DEATH | Specificity | 0.67<br>[0.60, 0.75] | 0.64 | 0.70 | 0.74 | 0.60 | 0.47 | 0.31 | 0.67 | 0.32 | 0.96 | 0.75 | 0.74 | 0.88 | 0.60 |
| ICU/DEATH | PPV | 0.73<br>[0.63, 0.82] | 0.71 | 0.80 | 0.75 | 0.73 | 0.70 | 0.70 | 0.82 | 0.63 | 0.55 | 0.90 | 0.71 | 0.74 | 0.54 |
| ICU | PPV | 0.67<br>[0.58, 0.77] | 0.68 | 0.75 | 0.68 | 0.71 | 0.67 | 0.63 | 0.81 | 0.52 | n/a | 0.81 | 0.71 | 0.74 | 0.47 |
| DEATH | PPV | 0.24<br>[0.15, 0.33] | 0.25 | 0.29 | 0.32 | 0.16 | 0.29 | 0.24 | 0.20 | 0.19 | 0.55 | 0.45 | 0.06 | 0.03 | 0.25 |
| ICU/DEATH | NPV | 0.83<br>[0.75, 0.91] | 0.83 | 0.68 | 0.82 | 0.79 | 0.82 | 0.80 | 0.85 | 0.79 | 0.92 | 0.82 | 0.91 | 0.98 | 0.79 |
| ICU | NPV | 0.86<br>[0.79, 0.94] | 0.86 | 0.75 | 0.89 | 0.81 | 0.86 | 0.82 | 0.86 | 0.84 | n/a | 0.89 | 0.91 | 0.98 | 0.85 |
| DEATH | NPV | 0.97<br>[0.93, 1.02] | 0.95 | 0.89 | 0.92 | 0.97 | 0.96 | 0.93 | 0.97 | 0.95 | 0.92 | 0.92 | 1.00 | 1.00 | 0.93 |

**Figure A1.**
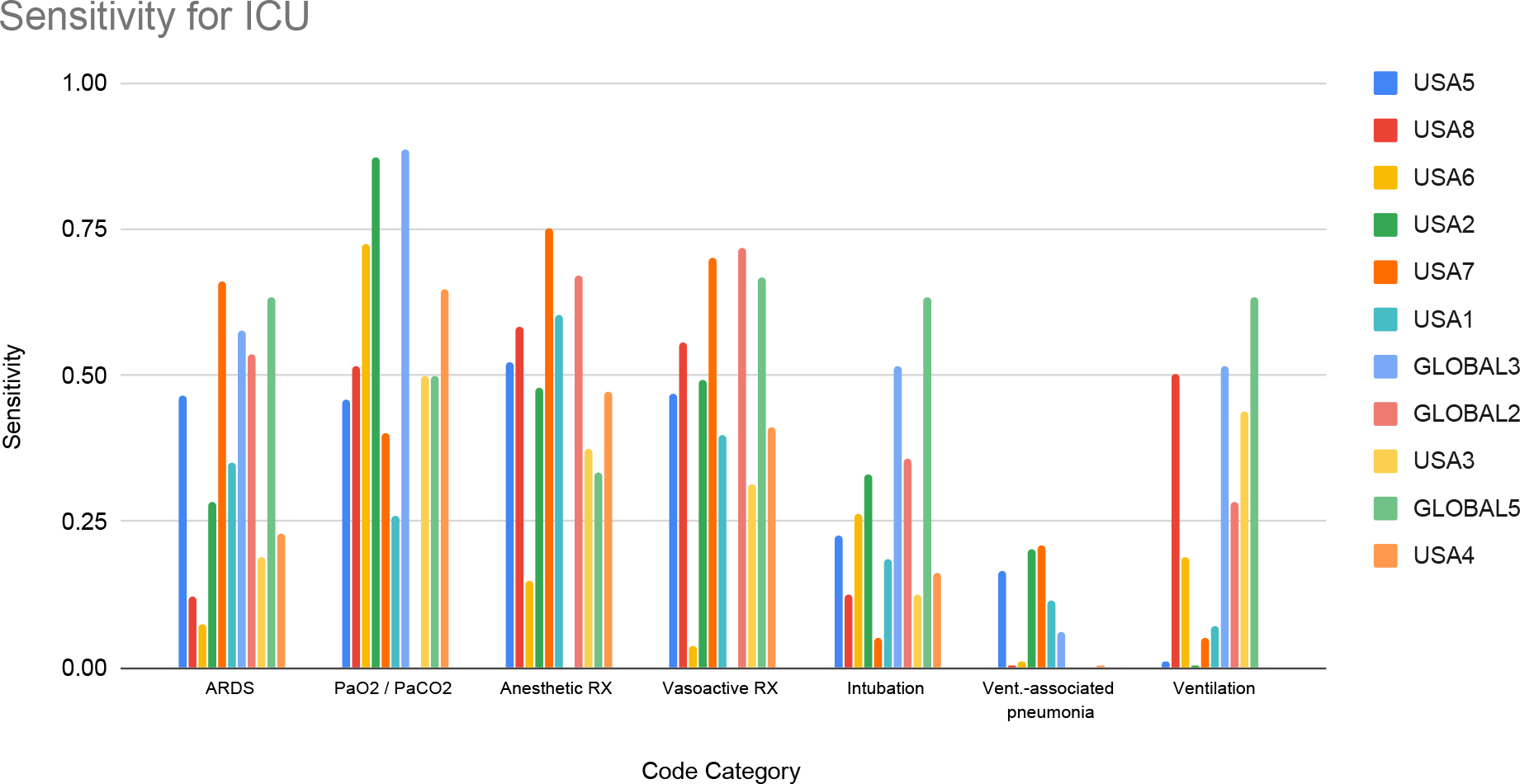
Sensitivity of code classes to identify ICU admission.

**Figure A2.**
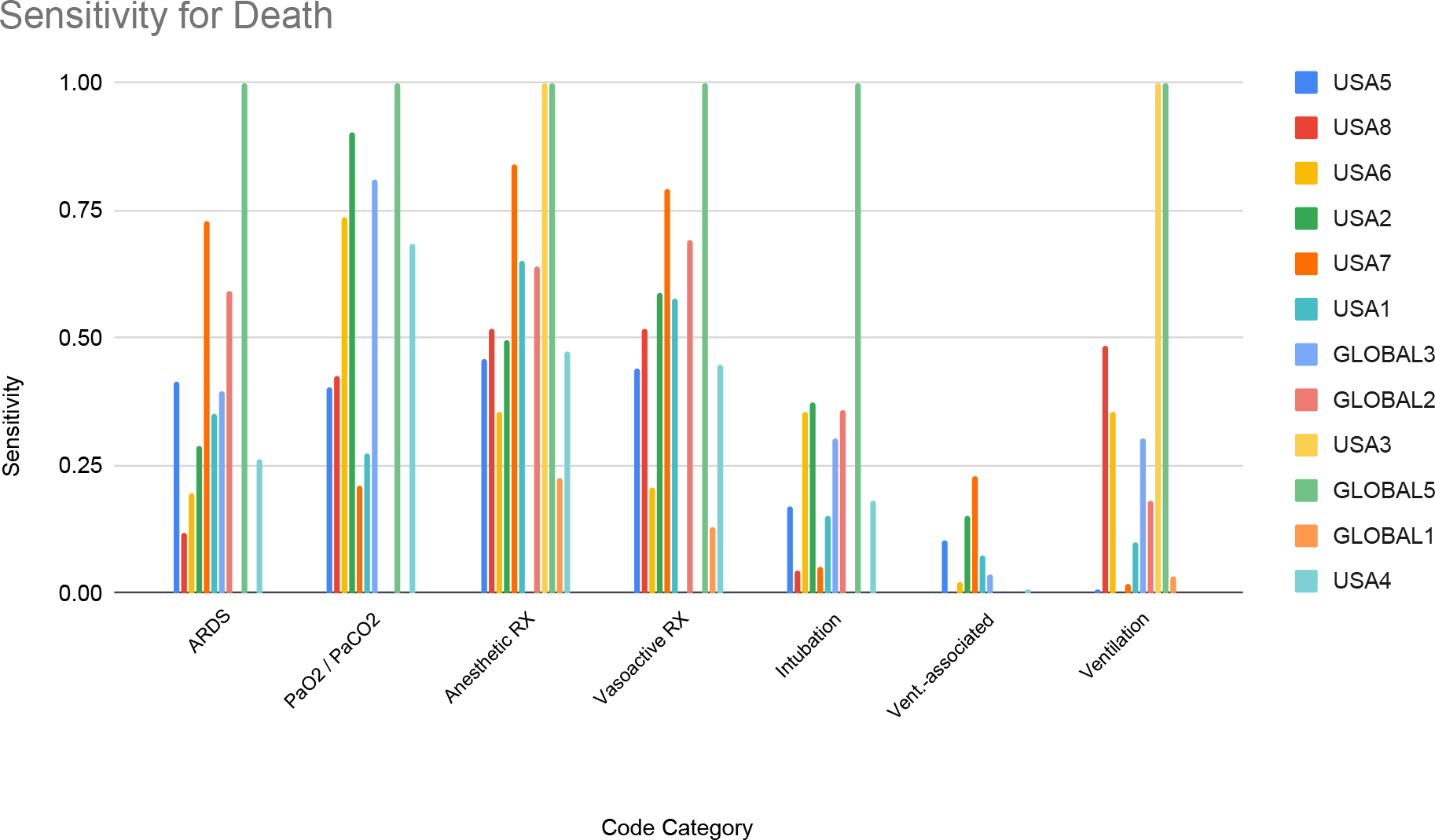
Sensitivity of code classes to identify death. (Sites close to 1.00 were biased by small populations.)

**Figure A3.**
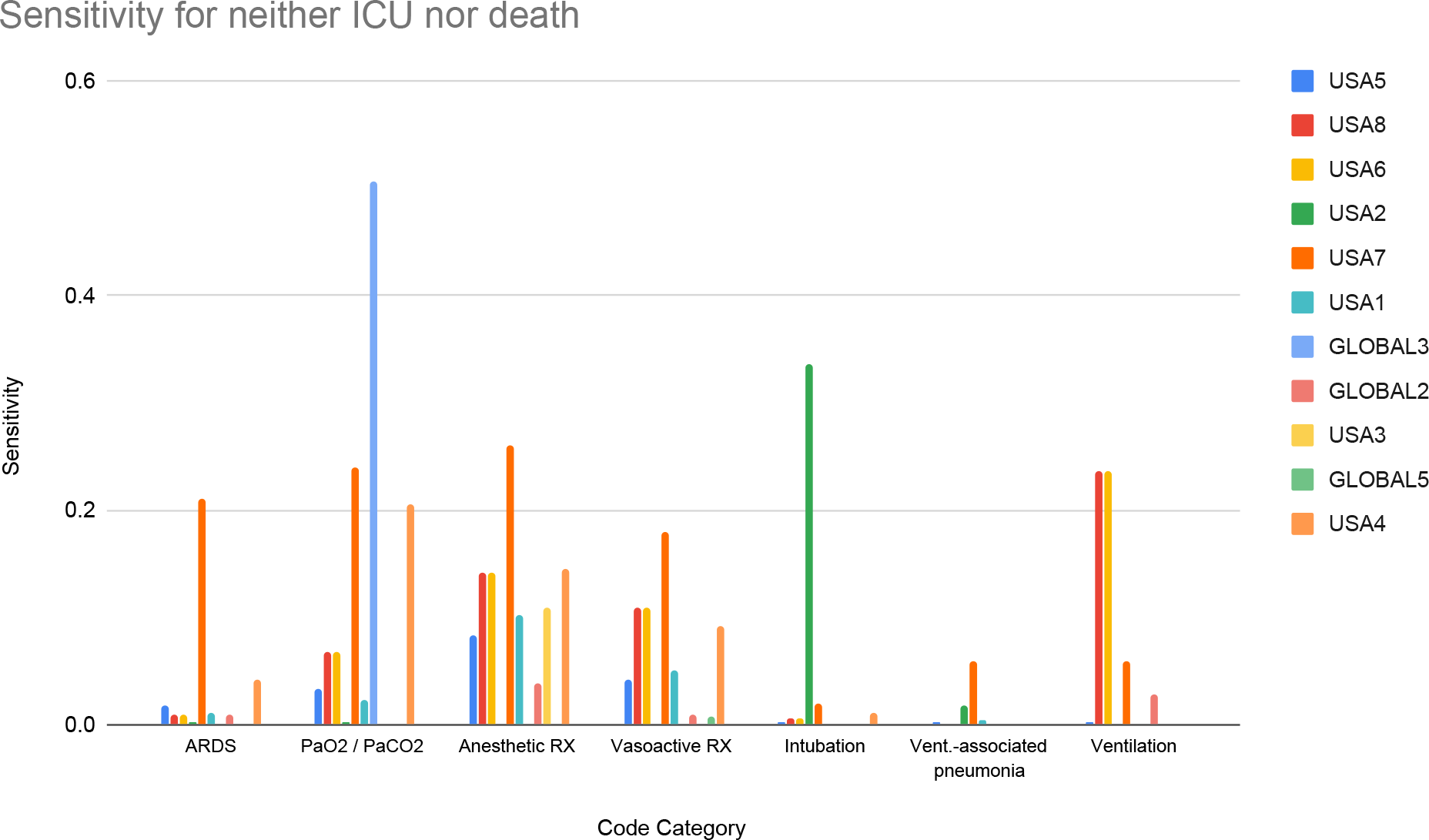
Sensitivity of code classes to identify no ICU admission nor death.

### Comparison of ICU Definitions

**Table A4.** Comparing chart-reviewed ICU admission data to other standards for finding ICU admission: hospital codes and CPT codes. This was done at Massachusetts General Hospital using the 4CE COVID-19 cohort and at UKFR using a manually chart reviewed subset of the 4CE COVID-19 cohort.

|  | MGH: CPT | MGH: Hospital | UKFR: Hospital |
| --- | --- | --- | --- |
| Sensitivity | 0.49 | 0.83 | 0.85 |
| Specificity | 0.99 | 0.99 | 0.88 |
| PPV | 0.49 | 0.97 | 0.78 |
| NPV | 0.59 | 0.92 | 0.93 |

## Appendix B: Additional Authorship Information

### Figure B1. 4CE Consortium Members

Adem Albayrak; Danilo F Amendola; Anthony L.L.J Li; Bruce J Aronow; Andrew Atz; Paul Avillach; Brett K Beaulieu-Jones; Douglas S Bell; Antonio Bellasi; Riccardo Bellazzi; Vincent Benoit; Michele Beraghi; José Luis Bernal Sobrino; Mélodie Bernaux; Romain Bey; Alvar Blanco Martínez; Martin Boeker; Clara-Lea Bonzel; John Booth; Silvano Bosari; Florence T Bourgeois; Robert L Bradford; Gabriel A Brat; Stéphane Bréant; Mauro Bucalo; Anita Burgun; Tianxi Cai; Mario Cannataro; Aize Cao; Charlotte Caucheteux; Julien Champ; Luca Chiovato; James J Cimino; Tiago K Colicchio; Sylvie Cormont; Sébastien Cossin; Jean Craig Juan Luis Cruz Bermúdez; Arianna Dagliati; Mohamad Daniar; Christel Daniel; Anahita Davoudi; Batsal Devkota; Julien Dubiel; Scott L DuVall; Loic Esteve; Robert W Follett; Paula S.A Gaiolla; Thomas Ganslandt; Noelia García Barrio; Nils Gehlenborg; Alon Geva; Tobias Gradinger; Alexandre Gramfort; Romain Griffier; Nicolas Griffon; Olivier Grisel; Alba Gutiérrez-Sacristán; David A Hanauer; Christian Haverkamp; Martin Hilka; John H Holmes; Chuan Hong; Petar Horki; Meghan R Hutch; Richard Issitt; Anne Sophie Jannot; Vianney Jouhet; Mark S Keller; Katie Kirchoff; Jeffrey G Klann; Isaac S Kohane; Ian D Krantz; Detlef Kraska; Ashok K Krishnamurthy; Sehi L’Yi; Trang T Le; Judith Leblanc; Guillaume Lemaitre; Leslie Lenert; Damien Leprovost; Molei Liu; Ne Hooi Will Loh; Yuan Luo; Kristine E Lynch; Sadiqa Mahmood; Sarah Maidlow; Alberto Malovini; Kenneth D Mandl; Chengsheng Mao; Patricia Martel; Aaron J Masino; Michael E Matheny; Thomas Maulhardt; Michael T McDuffie; Arthur Mensch; Bertrand Moal, Jason S Moore; Jeffrey S Morris; Michele Morris; Karyn L Moshal; Sajad Mousavi; Danielle L Mowery; Douglas A Murad; Shawn N Murphy; Kee Yuan Ngiam; Jihad Obeid; Marina P Okoshi; Karen L Olson; Gilbert S Omenn; Nina Orlova; Brian D Ostasiewski; Nathan P Palmer; Nicolas Paris; Lav P Patel; Miguel Pedrera Jimenez; Hans U Prokosch; Robson A Prudente; Rachel B Ramoni; Maryna Raskin; Siegbert Rieg; Gustavo Roig Dominguez; Elisa Salamanca; Malarkodi J Samayamuthu; Arnaud Sandrin; Emily Schriver; Juergen Schuettler; Luigia Scudeller; Neil Sebire; Pablo Serrano Balazote; Patricia Serre; Arnaud Serret-Larmande; Domenick Silvio; Piotr Sliz; Jiyeon Son; Andrew M South; Anastasia Spiridou; Amelia L.M. Tan; Bryce W.Q. Tan; Byorn W.L. Tan; Suzana E Tanni; Deanne M Taylor; Valentina Tibollo; Patric Tippmann; Andrew K Vallejos; Gael Varoquaux; Jill-Jênn Vie; Shyam Visweswaran; Kavishwar B Wagholikar; Lemuel R Waitman; Demian Wassermann; Griffin M Weber; Yuan William; Zongqi Xia; Alberto Zambelli; Aldo Carmona; Charles Sonday; James Balshi

**Table B1.** 4CE Authorship Contributions.

| Contribution | Conception or design of the work | Acquisition of data | Analysis or Interpretation of data | Drafting | Manuscript Approval |
| --- | --- | --- | --- | --- | --- |
| **Authors' Initials** | JK, GW, HE, PA, AM, GO, IK, GB, SM | JK, GW, HE, BM, PA, VC, TM, AM, AG, BB, AM, SV, GO, NY, KM, MB, KO, DM, MM, RF, DH, RB, JM, NL, DB, LC, VT, SR, AL, VJ, ES, MS, ZX, IK | JK, GW, HE, BM, PA, CH, VC, TM, AM, AG, BB, AM, AS, KM, MB, KO, DM, MM, RF, VT, SR, AL, VJ, ES, ZX, IK, SM | JK, HE, BM, PA, TM, AG, BB, AS, SV, KM, MB, NL, KW, ZX, IK, GB, SM | JK, GW, HE, BM, PA, CH, VC, TM, AM, AG, BB, AM, AS, SV, GO, NY, KM, MB, KO, DM, MM, RF, DH, RB, JM, NL, DB, KW, LC, VT, SR, AL, VJ, ES, MS, ZX, IK, GB, SM |

**Table B2.**
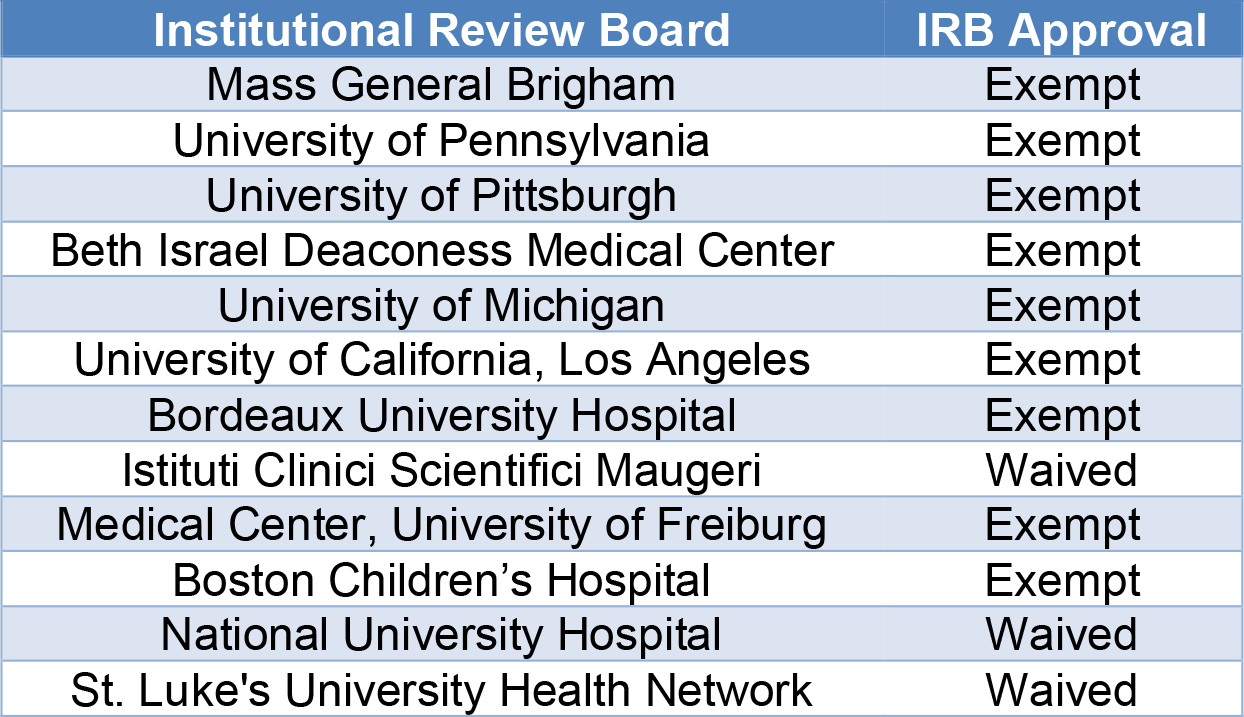
IRB Boards that Approved This Study.

| Institutional Review Board | IRB Approval |
| --- | --- |
| Mass General Brigham | Exempt |
| University of Pennsylvania | Exempt |
| University of Pittsburgh | Exempt |
| Beth Israel Deaconess Medical Center | Exempt |
| University of Michigan | Exempt |
| University of California, Los Angeles | Exempt |
| Bordeaux University Hospital | Exempt |
| Istituti Clinici Scientifici Maugeri | Waived |
| Medical Center, University of Freiburg | Exempt |
| Boston Children's Hospital | Exempt |
| National University Hospital | Waived |
| St. Luke's University Health Network | Waived |

